# ASSESSMENT OF KNOWLEDGE AND AWARENESS OF IRON DEFICIENCY ANEMIA AMONG PREGNANT WOMEN ATTENDING ANTE NATAL CLINIC AT FADA AND BAYAN FADA PRIMARY HEALTH CARE CENTRES IN BAUCHI METROPOLIS

**DOI:** 10.1101/2025.07.28.25332320

**Authors:** Muhammad Auwal Gidado, Mustapha Muhammad, Musa Bukar

**Affiliations:** Public Health Department, Jewel University, Gombe, Nigeria

**Keywords:** pregnant women, iron deficiency, anaemia

## Abstract

Anaemia is a condition in which blood does not contain enough haemoglobin that makes the blood to be red and also be able to carry oxygen that body cells need. In most developing countries of the world about 50 percent of all women, adolescent girls and children are anaemic. Anaemia in pregnancy is identified by the WHO as haemoglobin concentration less than 11g/dl. The commonest cause of anemia is iron deficiency. Iron is very important in the formation of haemoglobin. Iron deficiency anaemia is called nutritional anaemia. Nigeria is among countries with highest prevalence, about 52% are anaemic. The aim of this study is to determine the knowledge and awareness of iron deficiency anemia in pregnant women attending ANC at Fada and Bayan Fada Primary Health Care Centres in Bauchi metropolis. A quantitative cross-sectional study was employed to collect and analyzed all the result. Total number of 317 women were recruited using homogenous purposive sampling technique; their knowledge, dietary intake and socio-demographic information was obtained using a questionnaire. Out of 317 women, only 80 representing 25.2% of the total have adequate knowledge of Iron Deficiency Anaemia with average mean score of MS 3.70±2.532. The study confirmed a poor knowledge among women attending ante natal care at Fada and Bayan Fada Primary Health Care Centres, with lack of understanding of causes, effects, symptoms and preventive measures of Iron Deficiency Anaemia. It is recommended that primary prevention of increasing advocacy and awareness be prioritized by responsible Government ministries (health and education) and other relevant stakeholders (WHO, UNICEF and NCDC) in the country to increase knowledge of women attending Ante Natal Care towards Iron Deficiency Anaemia.

## INTRODUCTION

The term anemia is derived originally from the Greek word ‘an-haima’, which means “lack of blood”. Anaemia is a condition in which blood does not contain enough haemoglobin (haem-protein) that makes the blood to be red and also be able to carry oxygen that body cells need. Anaemia is found in all countries, rich and poor, and it affects women, elderly, and particularly children. In most developing countries of the world about 50 percent of all women, adolescent girls and children are anaemic (Jimoh, 2021). Peoples sometimes call anaemia “weak blood” or “thin blood” or “pale blood”; they may be because the person who suffer anemia may look pale or weak. The commonest cause of anemia is lack of nutrients iron or iron deficiency. Iron is very important in the formation of haemoglobin. Iron deficiency anaemia is a nutritional anaemia.

There are different types of anaemia common in human beings, these include: nutritional anaemia, sickle cell anemia, vitamin deficiency anaemia, anaemia of inflammation, aplastic anaemia, anaemia associated with bone marrow disease and haemolytic anaemia (Jimoh, 2021). Nutritional anaemia is a condition that occurs when the body cannot make enough haemoglobin and healthy red blood cells because it lacks the necessary nutrients particularly iron and proteins. Without adequate iron, the body can’t produce enough haemoglobin for red blood cells. Without iron supplementation, this type of anaemia occurs in many pregnant women. It is also caused by blood loss, such as from heavy menstrual bleeding, an ulcer, cancer and regular use of some over-the-counter pain relievers, especially aspirin, which can cause inflammation of the stomach lining resulting in blood loss. It occurs at all stages of the life cycle, but is more prevalent in pregnant women and young children. In 2002, iron deficiency anemia (IDA) was considered to be the most important contributing factors to the global burden of disease. Risk factors include a low intake and poor absorption of iron from diets high in phytate or phenolic compounds, and period of life when iron requirements are especially high (WHO, 2005) (i.e., growth and pregnancy).

Other risk factors such as heavy blood loss during menstruation and parasite infections such as hook worms, ascaris, and schistosomiasis, pregnancy, chronic conditions, family history, autoimmune disorders and alcoholism can lower blood hemoglobin (Hb) concentrations (Jimoh, 2021). Anemia in pregnancy is identified by the WHO as haemoglobin concentration less than 11g/dl and is divided into three levels of severity: mild anemia (Hb level, 9-10.9 g/dl), moderate level of anemia (Hb level, 7-8.9 g/dl) and severe anemia (Hb level 1-4.5 g/dl) (Margwe & Lupindu, 2018). Anemia in pregnancy is a major public health problem in developing countries. It is responsible for an estimated 20% of maternal deaths in West Africa and contributes to still more death through obstetric haemorrhage. Studies revealed a strong association between severe anemia and adverse perinatal and maternal outcome. Among the major causes of anemia in pregnancy are nutritional problems such as iron, folate and vitamin B12 deficiencies. Others include the adverse effects of malaria and hookworm infestations, infections such as human immunodeficiency virus (Dattijo, 2016).

## METHODOLOGY

### Description of the study area

The study was conducted in Bauchi Metropolis, Bauchi State. The study area has a total population of 670,000 as at 2023. Bauchi Metropolis is the capital of Bauchi state, Nigeria, which is located at 10° 19’ N, 9° 50’ E. The city lies on the Port Harcourt–Maiduguri Railway line. It consists of 8 administrative wards: Hardo ward, Dan Iya ward, Makama Sarkin Baki 1 ward, Makama Sarkin Baki 2 ward, Majidadi A ward, Majidadi B ward, Dawaki ward and Dankade ward.

#### Data Collection

Data collection was conducted over a period of two months (from 24th November, 2024 to 7th January, 2025) using a questionnaire as the main data collection tool. First and return visit women attending were systematically enrolled until the desired sample size was reached.

### Data Analysis

Data cleaning was done to all completed questionnaires soon after antenatal care (ANC) day each week by the principal investigator in collaboration with the assisting ANC staff in the clinic. A spreadsheet database was created using MS Excel 2010 for data entry. The completed data entry was imported to the Statistical Package for Social Science (SPSS) version 20.0 software for statistical analysis. Descriptive statistics were used to illustrate respondents’ socio-demographic characteristics. Categorical variables were measured as percentages while continuous variables were expressed as mean standard deviation. The Skewness and Kurtosis z-values, and Kolmogorov-Smirnov test were applied to declare the nature of data distribution. Inferential statistics (Mann-Whitney U Test and Kruskal-Wallis) were used to assess differences.

## RESULTS

Table 1 shows the socio-demographic characteristics of 317 pregnant women in the study area. The majority of the women are between the ages of 18–22 (39.7%). The majority of respondents were married (80.1%) and had secondary education (39.1%). Most of the women are housewives (49.5%), have 1 or 2 births (71.6%), and come from households with medium wealth (50.2%). The majority of the women are in their second trimester (39.7%). Almost 49.5% of the pregnant women were unemployed.

**Table 1:** Socio-demographic characteristics of the study participants (n=317)

| Characteristics | Frequency (n=317) | Percentage (%) |
| --- | --- | --- |
| **Age Group** |  |  |
| Below 18 years | 40 | 12.6 |
| 18-22 years | 126 | 39.7 |
| 23-27 years | 80 | 25.2 |
| 28-32 years | 48 | 15.1 |
| 33 years & Above | 21 | 6.6 |
| Undisclosed | 2 | 0.6 |
| **Marital Status** |  |  |
| Single | 34 | 10.7 |
| Married | 254 | 80.1 |
| Widow | 16 | 5.0 |
| Divorced | 8 | 2.5 |
| Undisclosed | 5 | 1.6 |
| **Educational Level** |  |  |
| None / Informal | 87 | 27.5 |
| Primary / Secondary | 182 | 57.4 |
| Tertiary / Undisclosed | 48 | 15.2 |

**Table 1 (Continued): Socio-demographic characteristics**
| Characteristics | Frequency (n=317) | Percentage (%) |
| --- | --- | --- |
| **Occupation** |  |  |
| Housewife | 157 | 49.5 |
| Civil servant | 48 | 15.1 |
| Business | 80 | 25.2 |
| None / Undisclosed | 32 | 10.1 |
| **Parity** |  |  |
| 0 birth | 37 | 11.7 |
| 1 birth | 108 | 34.1 |
| 2 births | 119 | 37.5 |
| 3+ births / Undisclosed | 53 | 16.8 |
| **Household Wealth Index** |  |  |
| Lowest | 112 | 35.3 |
| Medium | 159 | 50.2 |
| Highest / Undisclosed | 46 | 14.5 |
| **Trimester** |  |  |
| First Trimester | 75 | 23.7 |
| Second Trimester | 126 | 39.7 |
| Third Trimester / Undisclosed | 116 | 36.6 |

## DIETARY INTAKE DIVERSITY SUMMARY

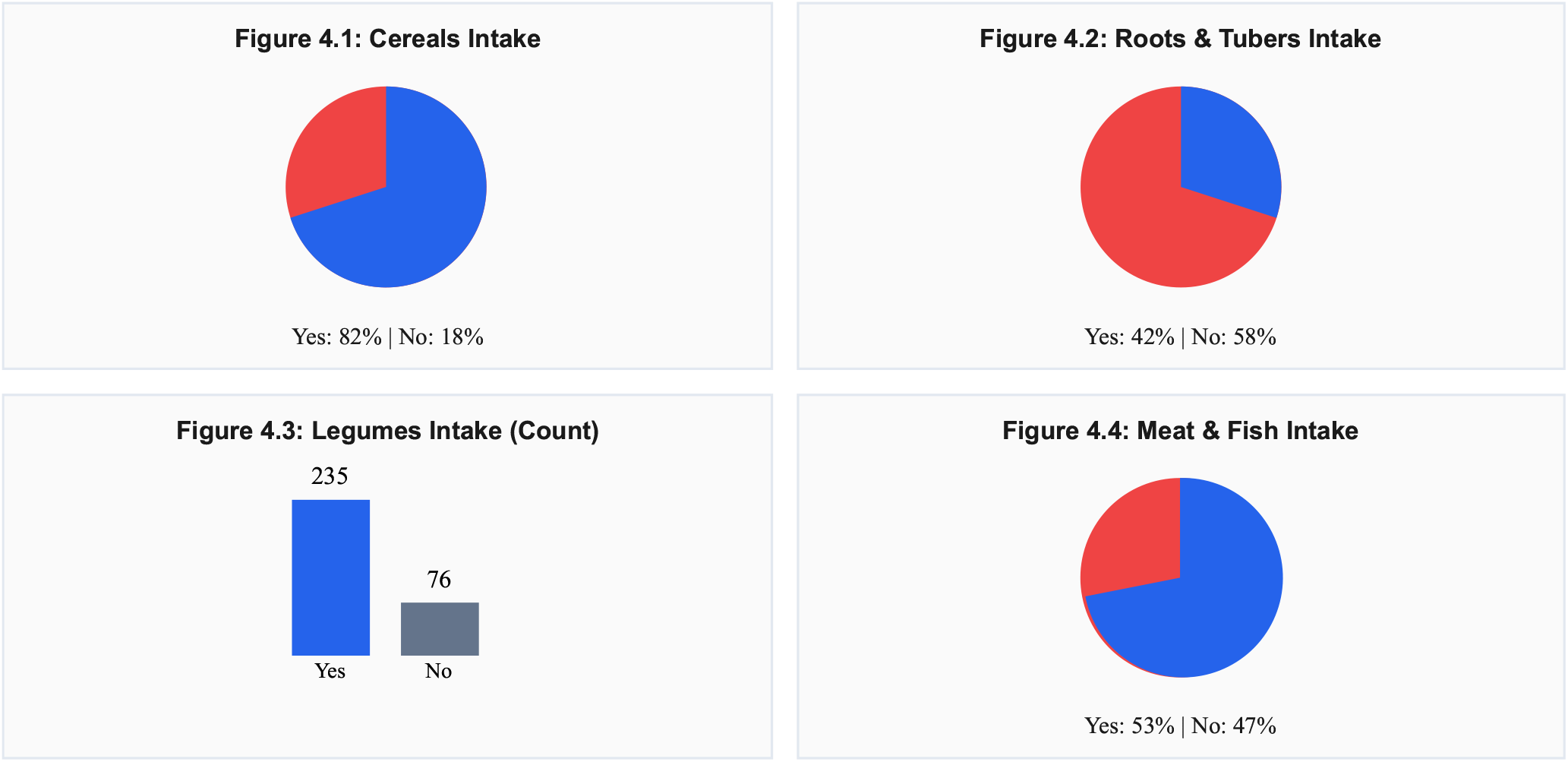
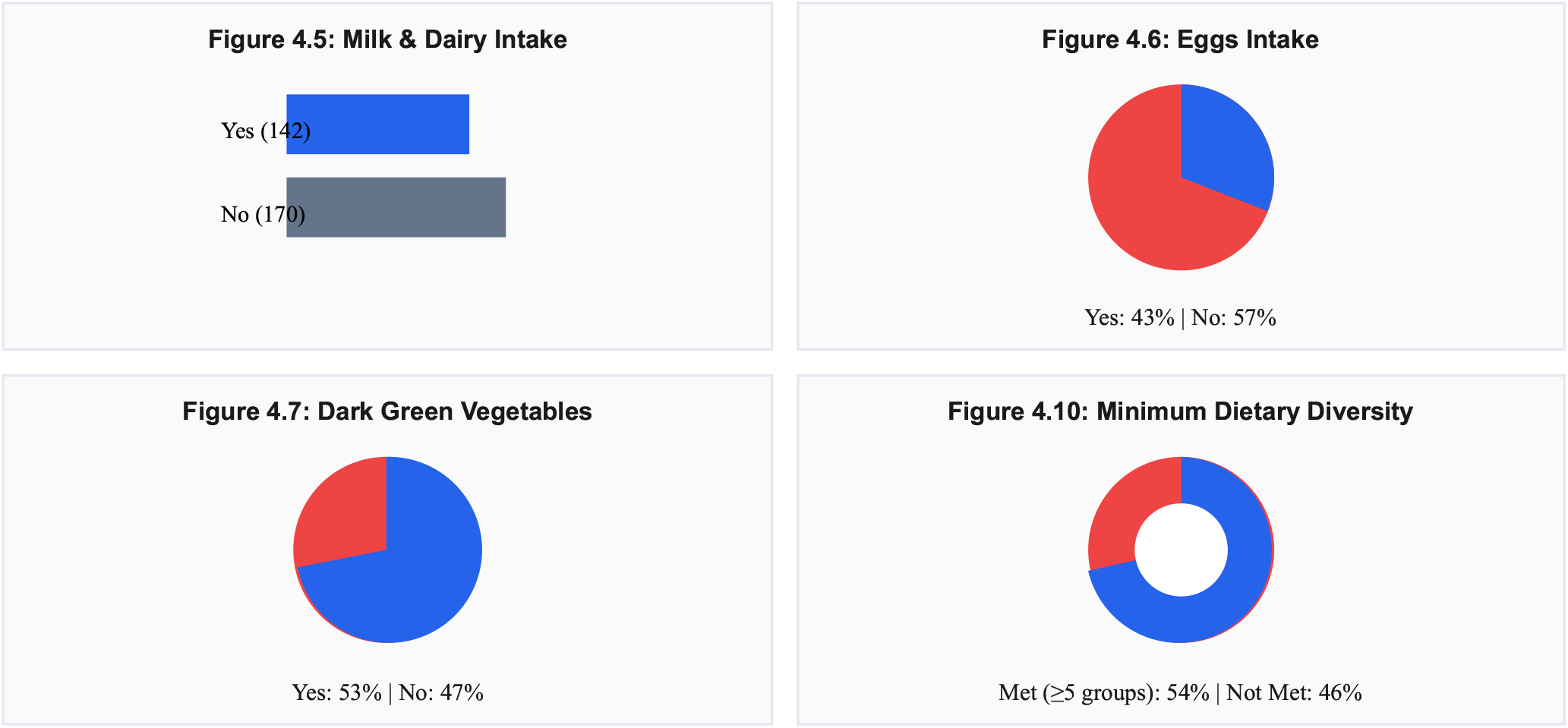

The dietary intake diversity scores of the study participants show that the most commonly consumed food groups are cereals (82.1%), legumes (74.1%), and milk and milk products (44.8%). The least commonly consumed food groups are eggs (42.3%), dark-green leafy vegetables (52.1%), and other vitamin A rich vegetables (45.4%). The percentage of individuals who met the minimum dietary diversity (consumed at least 5 out of 10 food groups) was 53.6%.

## ETHICAL APPROVAL

Ethical approval for this study was obtained from the Bauchi State Health Research Ethics Committee (HREC), located at the Ministry of Health, Bauchi State, Nigeria (Approval Reference No: ANC/ADM/0345, 12/10/2024). In addition, permission to conduct the study was obtained from Fada and Bayan Fada Primary Health Care Centres, which provided institutional support and access to the study site. The Ethics Committee confirmed that the study meets all ethical standards. All procedures performed involving human participants were in accordance with institutional and national ethical standards.

## CONCLUSION

In conclusion, the present study revealed a poor level of knowledge of Iron Deficiency Anaemia among pregnant women attending ante natal care at Bayan Fada and Fada Primary Health Care, Bauchi. Approximately 74.8% of the pregnant women attending Ante Natal Care in the current study had a low level of knowledge towards Iron Deficiency Anaemia with a lack of understanding of the causes, effects, symptoms, and preventive measures including iron-rich dietary intake. Extensive awareness and effective advocacy models for behavior change should be delivered for pregnant women attending Ante Natal Care via health facilities and print media in sub-Saharan and rural settings.

## Data Availability

All data produced in the present work are contained in the manuscript

## RECOMMENDATIONS

1. Primary prevention through increased advocacy and awareness of Iron Deficiency Anaemia should be prioritized by responsible government ministries (health, education, and social welfare) and relevant stakeholders (WHO, UNICEF, NCDC) to improve knowledge levels among women attending Ante Natal Care.
2. Findings can be used to inform interventions and policies aiming to improve the health and well-being of pregnant women and their babies.
3. Further studies should focus on larger populations and investigate the relationship between dietary intake diversity and knowledge of iron deficiency anaemia among pregnant women.

